# Linking School Stress and Psychosomatic Complaints in South Tyrol, Northern Italy: Parental and adolescents’ perspectives in a cross-sectional design

**DOI:** 10.64898/2026.05.26.26354140

**Authors:** Verena Barbieri, Giuliano Piccoliori, Adolf Engl, Doris Hager von Strobele Prainsack, Christian J. Wiedermann

**Author notes:** Correspondence, Verena Barbieri, PhD, Institute of General Practice and Public Health Claudiana—College of Health Professions Lorenz Böhler Street 13, 39100 Bolzano, Italy.

## Abstract

**Background:** School stress and psychosomatic complaints are linked and increase in high-income countries, with differences between countries. Evidence of how these parameters develop in Italy, particularly through combined parental and self-reported perspectives across age and gender, is limited.

**Methods:** A population-based online survey investigated school stress and psychosomatic complaints in children and adolescents aged 6–19 years, analyzing proxy- and self-reported data based on standardized validated instruments. Data was stratified by gender and age for children (6–10), early adolescents (11–14), and late adolescents (15–19).

**Results:** For early and late adolescents, the gender gap was evident, with higher levels of stress and health complaints in late adolescent girls. In this group, 56% of the girls self-reported rather/high school stress, and 43% of the boys. Parents perceived school stress and psychosomatic problems of their children as less severe than adolescents themselves. Parents stated a higher effect of parental help with school problems, and a lower effect of physical activity and digital media use on their children’s psychosomatic problems. Physical activity was related to fewer psychosomatic complaints, especially in girls.

**Conclusions:** This study identified late adolescent girls as vulnerable group, underscoring the critical need for gender-specific early prevention strategies starting in childhood, particularly for families with lower socioeconomic status. Parental perspectives may underestimate adolescents’ stress levels and psychosomatic well-being. In early adolescence, less digital media use may prevent psychosomatic problems, in late adolescence, physical activity may be a preventive method. Further longitudinal investigations should put a special focus on self-and proxy-reported perspectives.

## 1. Introduction

Over the past three decades, a consistent association between school-related stress and psychosomatic complaints in children and adolescents has been documented across multiple European countries (1),(2),(3),(4). Psychosomatic complaints, including headaches, stomachaches, sleep problems, and irritability, are among the most common health problems reported by school-aged populations, and schoolwork pressure has been identified as one of their principal correlates (2), (4). Cross-national data from the Health Behavior in School-aged Children (HBSC) consortium indicate that both psychosomatic complaints and perceived schoolwork pressure have increased over time in high-income countries, although with substantial variation between nations (2).

In addition to the direct association between school stress and psychosomatic complaints, several factors have been identified as potential correlates. Social support from family and peers (1),(5) and socioeconomic status (6) appear to play protective roles. Physical activity is inversely related to both school stress and psychosomatic complaints (3), and recently, screen time and digital media use have attracted attention as correlates of psychosomatic complaints (7). Emerging evidence suggests that managing digital information for school tasks presents distinct challenges for younger adolescents (8).

Gender disparities in psychosomatic health have been extensively studied. Girls report more psychosomatic complaints than boys, a disparity that has widened over the last decades (9),(10). This pattern has been attributed to the “educational stressors hypothesis,” which posits that modern societies’ emphasis on educational achievement generates increased performance expectations, testing, and evaluation pressures, and that girls — who tend to place greater value on academic success and whose self-esteem is more closely tied to academic performance — are disproportionately affected (11),(12). Longitudinal HBSC data spanning 1993 to 2017 indicated that school stress accounts for a substantial portion of the increase in symptoms for girls but only a minor share of the increase for boys (13). The authors found weak evidence for the educational stressors’ hypothesis regarding the overall trend in symptoms but strong evidence for it in explaining the growing gender gap. Girls also report considerably higher levels of mental health problems toward the end of compulsory schooling (14).

Evidence shows that frequency, number, and persistence of psychosomatic complaints during adolescence were associated with symptoms of mental health problems in young adulthood (15),(16),(17),(18),(19). Thus, early attention, monitoring, and preventive interventions are important. In this context, the role of parents in dealing with psychosomatic complaints and school stress needs attention. While evidence about mental health problems shows that parental and adolescent perceptions are often different (20),(21),(22), evidence about perceived psychosomatic health complaints and school stress among adolescents is lacking. Prior research has demonstrated that parent and child reports of psychosomatic complaints and mental health can diverge considerably (23) and that relying solely on proxy reports may result in affected adolescents being overlooked (24),(25).

Despite this considerable body of international evidence, country-level differences in both the prevalence and trends of psychosomatic complaints and school stress require caution against cross-national generalization (2). In Italy, the HBSC data indicate that Italian adolescents display symptom patterns comparable to European averages, with an increasing trend over the last two decades (26),(27), where females and lower family affluence were associated with more psychosomatic symptoms. During the pandemic, Italian adolescents exhibited higher rates of mental health issues (28). A comprehensive understanding of how school stress, psychosomatic complaints, and gender differences interact across different developmental stages post-pandemic, from childhood through late adolescence, requires further investigation.

In South Tyrol, a region in the North of Italy at the border to Austria and Switzerland, four repeated cross-sectional population-based surveys (COP-S: Corona and Psyche South Tyrol) among children and adolescents between 6 and 19 years have been conducted since 2021, screening for mental health disorders and psychosomatic complaints (29),(30). Preliminary results indicate that mental health outcomes and psychosomatic symptoms have not reverted to pre-pandemic levels, highlighting the need for targeted prevention and intervention strategies to address these issues.

Importantly, the study design includes both parental proxy reports (for children aged 6–19 years) and adolescent self-reports (for those aged 11–19 years), which is of particular methodological relevance, as no Italian study has systematically compared parental and self-reported perspectives on school stress and psychosomatic complaints across multiple age groups to date.

Building on the most recent COP-S wave, this study is the first to integrate population-based dual-informant data on school stress, psychosomatic complaints, and associated factors across three developmental stages in a post-pandemic Italian cohort. Using standardized and validated assessment instruments, this cross-sectional study aimed to examine:

This study aims to explore school stress and psychosomatic complaints

1. From parental and adolescents’ perspectives stratified by gender and age
2. Discrepancies in parental and adolescent’s perceived school stress and psychosomatic complaints
3. Associations of school stress and psychosomatic complaints with lifestyle factors from a dual perspective

By combining parental and self-reported data across a large regional cohort and three developmental stages, this study aims to elucidate the complex interactions between school stress, psychosomatic complaints, and social and lifestyle factors, and to generate evidence for prevention.

## 2. Methods

A population-based online survey was conducted between 17 March and 13 April 2025 using the SoSci Survey platform (Version 3.2.46, Munich, Germany) for the fourth time. This design, employing both proxy-reported data for younger children (6–19 years) and self-reported data for adolescents (11–19 years), was specifically chosen to capture divergent perspectives on school stress and psychosomatic complaints across different developmental stages and to allow for cross-informant comparisons that are crucial to our research questions. Recruitment was carried out through all provincial schools by contacting parents via email and sending a link to the anonymous questionnaire. A reminder email was sent two weeks later. Informed consent was obtained from the parents and adolescents. More than 40,000 families were invited to participate in the study. Parents first completed the proxy version before the adolescents filled in the self-report form.

### 2.1. Assessment of parental and adolescents perceived school stress

School-related stress was assessed using the question, “How much does your child feel stressed by schoolwork?”, ”How stressed do you feel because of your schoolwork? with possible answers ranging from ‘Not at all’ (0) to ‘Very’ (3). This single-item measure of perceived schoolwork pressure was derived from the HBSC study protocol, where it has been employed across multiple survey cycles in over 40 countries (31). The item has demonstrated adequate criterion validity in large-scale epidemiological surveys and is considered an established indicator of school-related stress in adolescent health research (11).

### 2.2. Sociodemographic variables

The collected sociodemographic variables included children’s age and gender, parental education as per the Comparative Analysis of Social Mobility in Industrial Nations (CASMIN) index (32), single parenthood, and migration background.

Perceived social support was measured with Multidimensional Scale of Perceived Social Support (MSPSS) (33), and family socioeconomic status with the Family Affluence Scale III (FAS III) (34),(35),(36),(37).

All sociodemographic variables were retrieved from proxy reports.

#### 2.2.4. Lifestyle factors

Parental help with school issues was assessed using the question, “How often have you assisted your child with schoolwork in the current school year?”, “How often have your parents assisted you with schoolwork in the current school year?”, with answers on a 5 point Likert scale from 1=”never” to 5=”always.”

Digital media use for school and private issues was assessed with the question: “How many hours does your child actually spend with digital media (Computer, Smartphone, Tablet…) for school/private concerns?”, “How many hours do you actually spend with digital media (Computer, Smartphone, Tablet…) for school/private concerns?” answers ranged from1=”none” to 7=”five or more hours.”

Physical activity was assessed with the question: “Over the past week, on how many days did your child engage in at least 60 minutes of sports or physical activity?” “Over the past week, on how many days did you engage in at least 60 minutes of sports or physical activity?” Answers were available on a scale ranging from 1 (0 days) to 8 (7 days).

#### 2.2.3. Psychosomatic complaints

The Health Behavior in School-aged Children Symptom Checklist (HBSC-SCL) was used to assess psychosomatic complaints. This checklist identifies psychosomatic complaints with items such as, “How often has your child had headaches in the past week?” Eight psychosomatic problems were assessed: headaches, stomachaches, backaches, feeling down, irritability, feeling nervous, sleep problems, and dizziness. Responses were recorded on a 5-point scale ranging from 1 = “daily” to 5 = “not at all” (38),(31).The total score ranged from 8–40. For the analysis, the sum score was inverted to obtain higher values for more complaints.

### 2.3. Data Analysis

For separate proxy- and self-reported analyses, we analyzed all datasets in which psychosomatic complaints and school stress were reported completely by the corresponding participant. For dual perspective analyses, proxy and self-reports, where parents and children provided complete answers, were analyzed.

Descriptive statistics of nominal or categorical variables were presented as absolute counts and percentages, and distributions were visualized as bar plots. Metric data were presented as means (m) ± standard deviations (SD), and distributions of metric variables were visualized using box plots. Lifestyle parameters were dichotomized for the descriptive statistics.

Group differences were tested using chi-square tests for nominal and categorical variables, Mann–Whitney tests for non-normally distributed continuous variables to compare two groups, and Kruskal–Wallis tests to compare three or more groups. Pairwise post hoc testing was performed using the Bonferroni correction.

Paired metric variables of proxy- and self-reported data were analyzed using paired sample Wilcoxon tests and one-way random effects intraclass correlation (ICC) for absolute agreement of mean ratings for two different raters (39). Values <0.5 indicated poor reliability, values between 0.5 and 0.75 indicated moderate reliability, values between 0.75 and 0.9 indicated good reliability, and values > 0.9 indicated excellent reliability.

Paired categorical variables of proxy- and self-reported data were analyzed using paired Wilcoxon tests and Cohen’s kappa. Values <0.4 indicated slight, values from 0.4 to 0.6 moderate and values from 0.6-0.8 substantial and values >0.8 excellent agreement.

Associations between dichotomous and continuous variables we used Point-Biserial correlation and for ordinal and continuous variables Spearman’s correlation coefficient.

The reliability of the multi-item scales was assessed using Cronbach’s alpha.

p–values < 0.001 are indicated with ***, < 0.01 with **, < 0.05, *, and p-values ≥ 0.05 are considered non-significant (n.s.). All statistical analyses were performed using SPSS version 27.0.0.0.

## 3. Results

The number of participating parents was 7,818, of which 5,820 had complete datasets regarding proxy-reported adolescents’ psychosomatic complaints and school stress. In the age group from 6 to 10 years, we could evaluate 2,532 cases, in the group from 11 to 14 years, 1,699 cases, and in the group from 15 to 19 years, 1,589 cases. Of the 2,554 participating adolescents, there were 1,564 evaluable self-reports, 818 in the 11–14 age group and 746 in the 15–19 age group.

In the following chapters, we compared variables between the three age groups and gender and analyzed the main outcomes and their associations with mental health.

### 3.1. Demographic characteristics and dichotomized lifestyle parameters

Table 1 shows the demographic characteristics of the participants in all three age groups. Gender of children did not differ significantly between the three age groups. The percentage of single parenthood increased significantly with increasing children’s age, and the percentage of participants with a migration background increased significantly with decreasing children’s age. FAS III was significantly lower in younger age groups and higher in older ones. Parental education levels were significantly higher for younger children. The hours spent on sports decreased with increasing age. MSPSS differed significantly between age groups, with higher levels found more often in younger children and medium levels in older children. More than two hours of digital media use for school and private concerns was reported significantly more often by parents of older children. Parental help with school problems was reported more frequently by parents of children and young adolescents. The percentage of parents reporting three hours or more of physical activity of their child decreased with increasing age of the children.

**Table 1.**
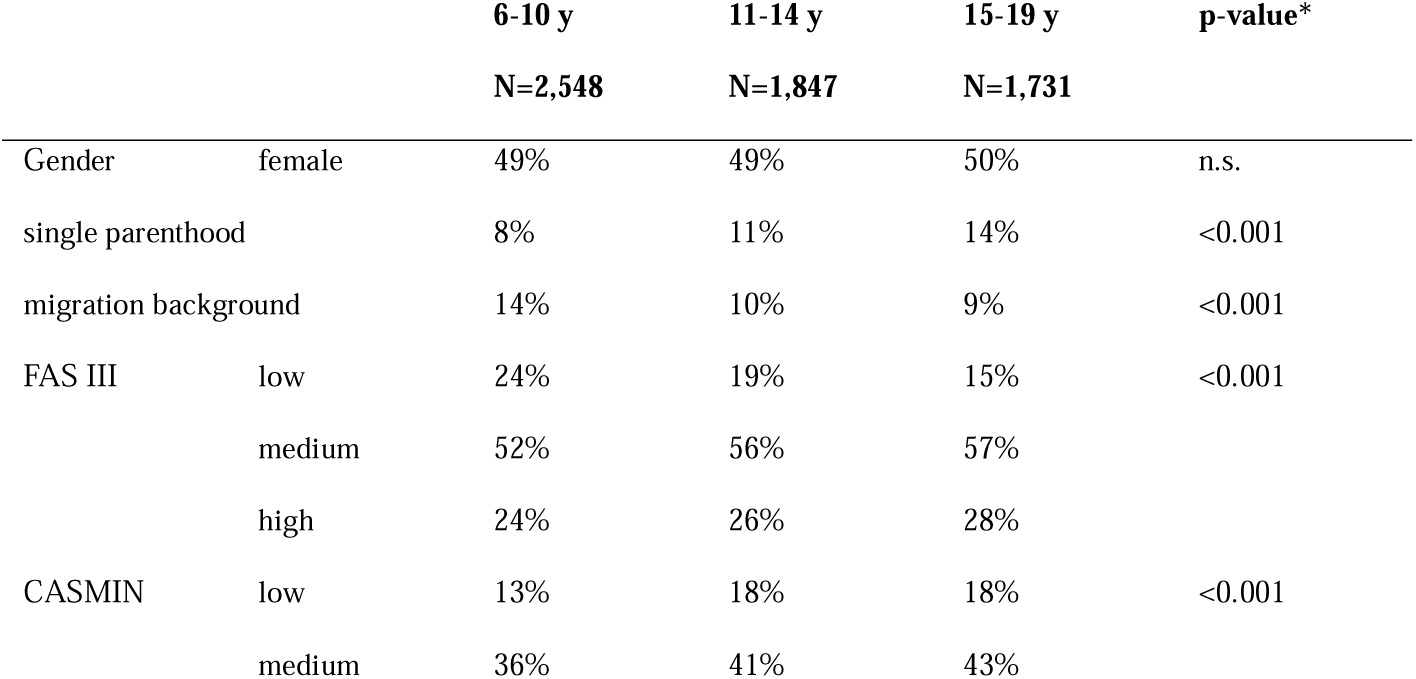
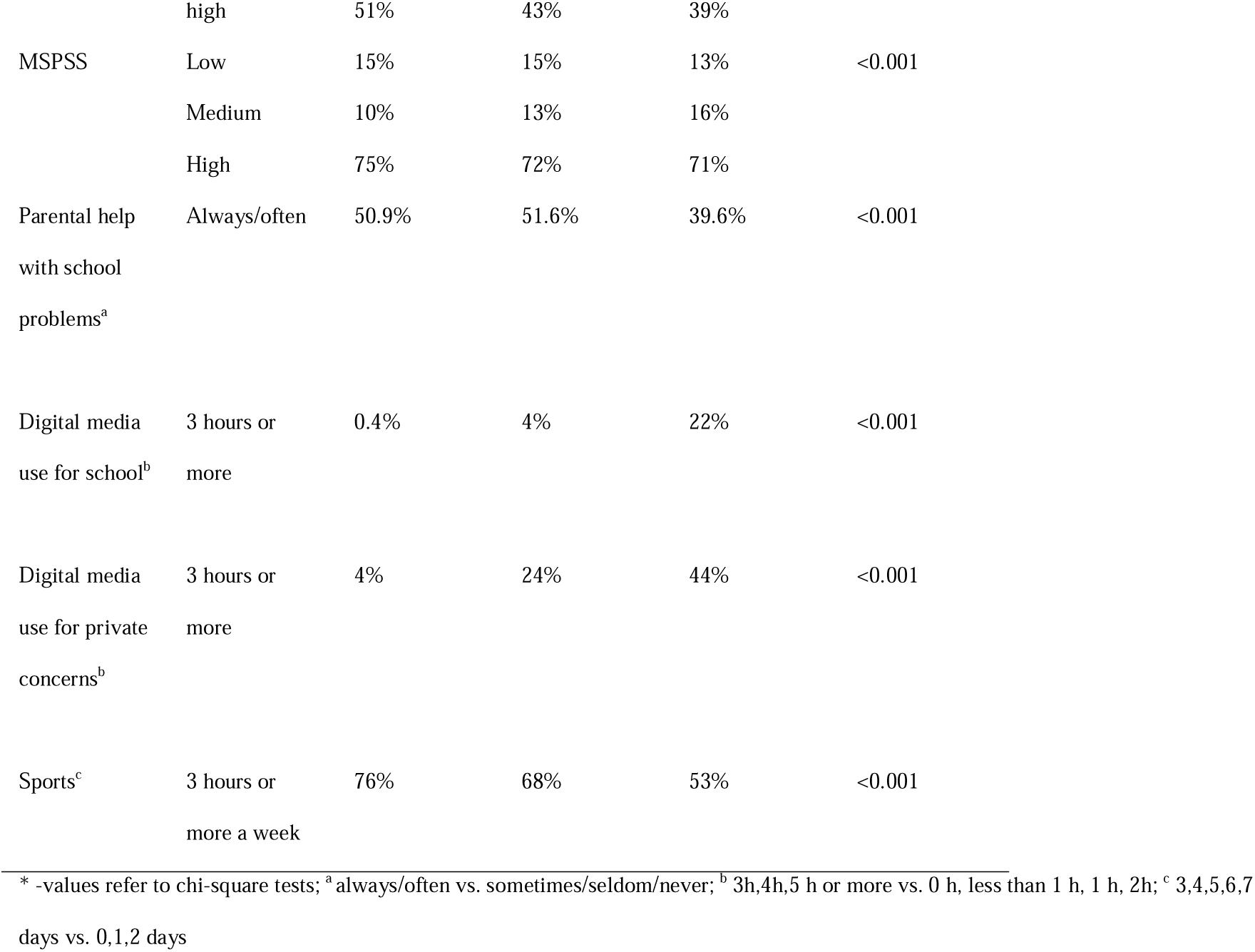
Demographic situation for the age groups from 6 to 10, 11 to 14 and 15 to 19 years (proxy-reports)

### 3.2. Gender disparities and relations to demographic variables

Table 2 presents the effect sizes of gender for school stress, psychosomatic complaints, and lifestyle per age group. In the age group from 6 to 10, small effect sizes with higher school stress, more parental help with school issues, and higher digital media use for private concerns for boys were found. Boys had more physical activity per week than girls. In the age group from 11 to 14, significant effect sizes were found for psychosomatic complaints and hours of digital media use for school concerns in proxy-reports, with females having higher values, while boys reported significantly more physical activity. The same pattern for psychosomatic complaints and physical activity was found in the self-reports. In the age group from 15 to 19, proxy and self-reports showed higher school stress, higher use of digital media for school issues, and higher psychosomatic complaints for girls, and more digital media use for private concerns and more physical activity for boys.

**Table 2.** Point biserial coefficient for female gender with psychosomatic complaints, school stress and lifestyle parameters.

|  | proxy |  |  | self |  |
| --- | --- | --- | --- | --- | --- |
|  | 6-10 years | 11-14 years | 15-19 years | 11-14 years | 15-19 years |
| **School stress** | -0.048* | n.s. | 0.114*** | n.s. | 0.169*** |
| **Sum score of psychosomatic complaints** | n.s. | 0.164*** | 0.159*** | 0.216*** | 0.212*** |
| **Parental help** | -0.058** | n.s. | n.s. | n.s. | n.s. |
| **Hours of digital media use (school)** | n.s. | 0.095*** | 0.180*** | n.s. | 0.164*** |
| **Hours of digital media use (private)** | -0.070*** | n.s. | -0.101*** | n.s. | -0.101*** |
| **Physical activity** | -0.192*** | -0.217*** | -0.211*** | -0.197*** | -0.215*** |

Differences in self- and proxy-reported outcomes regarding demographic variables were tested and are reported with Bonferroni correction for the seven demographic variables in each age group (significance level p=0.05/5=0.0071).

Self-reported school stress was significantly higher in the 11–14 age group for single parenthood (p=0.006).

In the 6–10-year age group, proxy-reported school stress was significantly higher for lower CASMIN index (p<0.001) and lower FAS III (p<0.001), and it differed for MSPSS (p=0.001), with the highest values in the moderate group. In the 11–14 age group 11 14 higher levels were found for single parenthood (p=0.002) and a lower CASMIN index (p=0.002). In the 15–19 age group, higher levels of school stress were found for single parenthood (p=0.001) and moderate MSPSS (p<0.001).

Proxy-reported sum scores of psychosomatic complaints were higher for single parenthood in the 6–10 (p < 0.001) and 15–19 (p < 0.001) age groups. For the MSPSS (Figure 1), there was a significant difference in the age groups 6–10 (p<0.001, see figure 3), 11–14 (p<0.001), and 15–19 (p<0.001).

**Figure 1.**
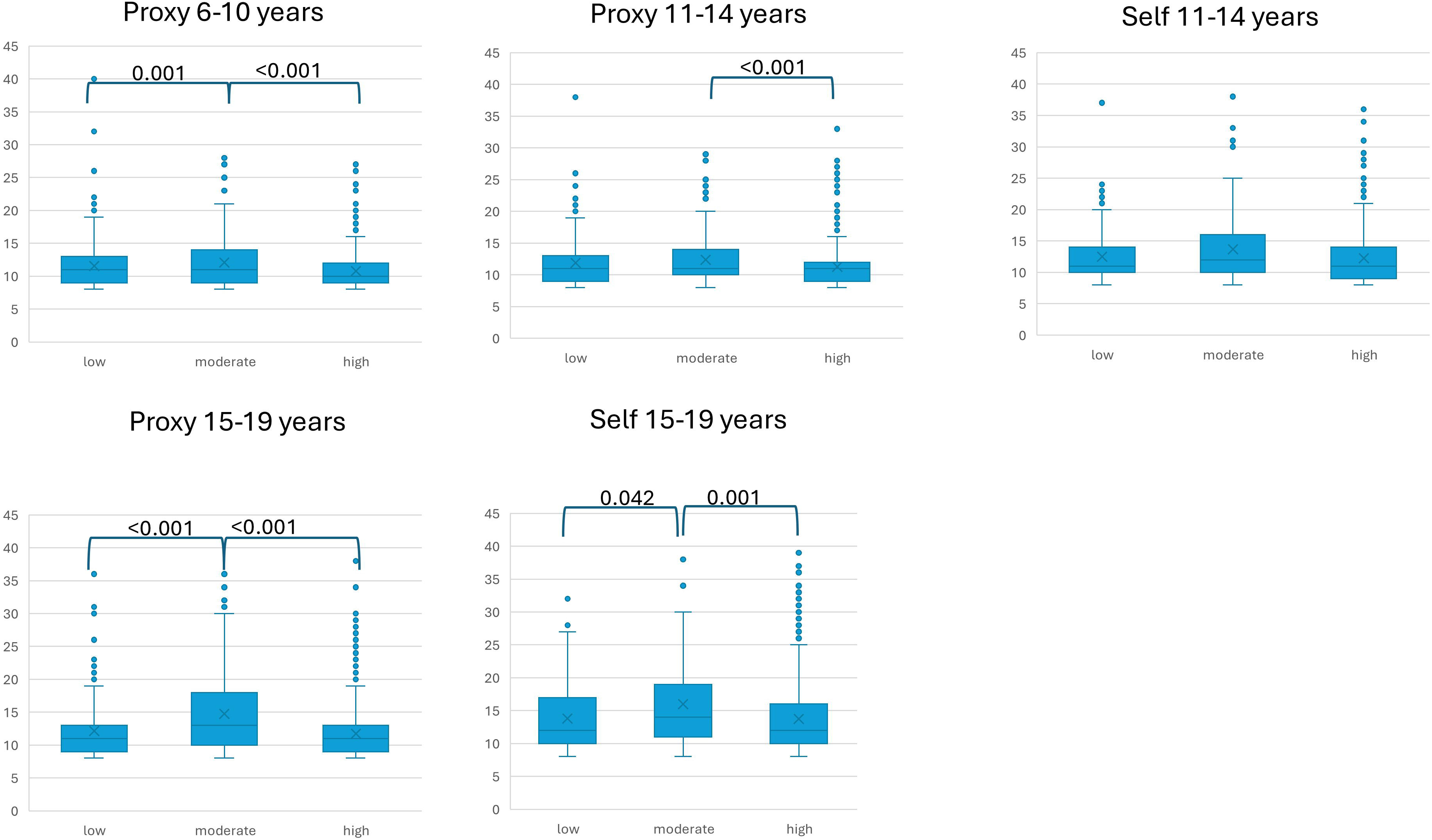
shows the significant differences in the sum scores of psychosomatic complaints per MSPSS category. Upper panel: proxy-reports for children and proxy- and self-reports for early adolescents; lower panel: proxy- and self-reports for late adolescents

The self-reported number of psychosomatic complaints was significantly higher for single parenthood in the 11–14 age group (p=0.004). For the MSPSS (Figure 1), there was a significant difference in the age group from 15 to 19 (p=0.001).

### 3.3. A dual perspective: Proxy- and self-reported school stress and psychosomatic complaints

Detailed percentages per age group for males and females of proxy- and self-reported main outcomes are provided in Supplement S1.

Proxy-reported school stress (Figure 2) differed significantly between age groups for boys (p=0.006) and girls (p<0.001), with higher levels of stress in older participants. Self-reported school stress differed between 11-14 aged and 15-19 aged boys (p<0.001) and girls (p<0.001) significantly with higher stress in the older age group. Proxy- and self-reported school stress differed significantly only in girls aged 15-19 years (p=0.001) with higher levels of school stress in self-reports.

**Figure 2.**
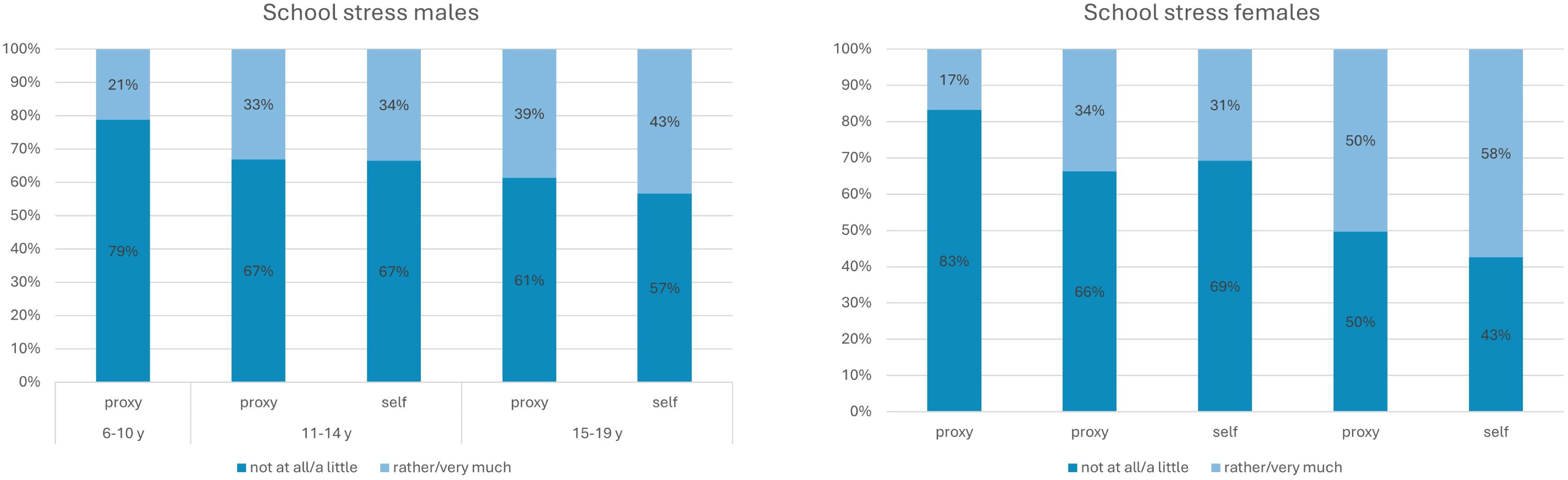
Proxy- and self-reported school stress for males and females per age group

For proxy-reported sum scores of psychosomatic complaints (Figure 3) a significant difference between age groups was found for girls (p<0.001) with higher scores in older groups. For self-reported scores, the difference was significant for boys (p<0.001) and girls (p<0.001) with higher scores in the older age group. The sum score of psychosomatic complaints was significantly higher in self-reports than in proxy-reports for girls aged 11–14 (p<0.001) and 15–19 (p<0.001) and for boys aged 11–14 (p=0.001) and 15–19 (p<0.001).

**Figure 3.**
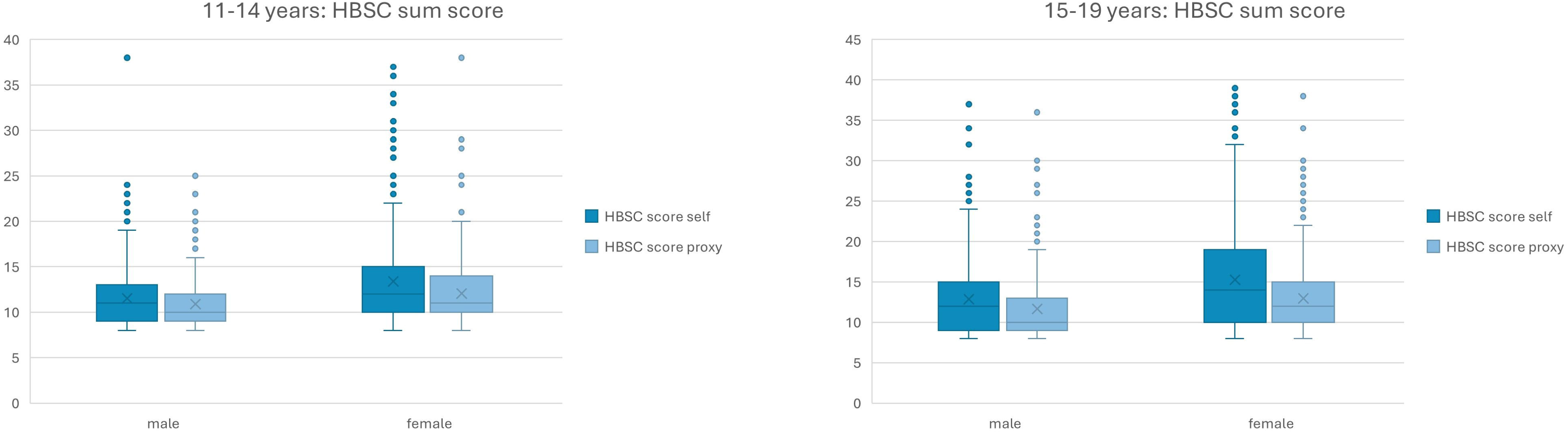
Proxy- and self-reported percentages of psychosomatic complaints in terms of HBSC-SCL sum scores as box plots stratified per age group and gender

Reliability analyses returned a moderate effect between proxy- and self-reported data for school stress and HBSC scores (Table 3). For school stress (Cohen’s Kappa) and psychosomatic complaints (ICC), younger males showed the lowest agreement between proxy and self-reported data. The percentages of agreement and disagreement in terms of </> are listed in Table 3, while the absolute differences between proxy-rated school stress and – self-rated school stress) are presented in Figure 4. Self-rated school stress was found to be slightly higher than parent-rated school stress, especially among girls aged 15–19. The means with 95% confidence intervals and 5% trimmed means of the HBSC sum score differences (proxy-self) are shown in Table 3. Negative differences indicated higher sum scores in self-reports for both genders and age groups, with the highest differences for girls aged 15–19 years. The box plots of the HBSC sum score differences (proxy-self) are presented in Figure 5, showing the largest negative median differences for girls from 15 to 19 and nearly 0 distributed differences for boys from 11 to 14.

**Figure 4.**
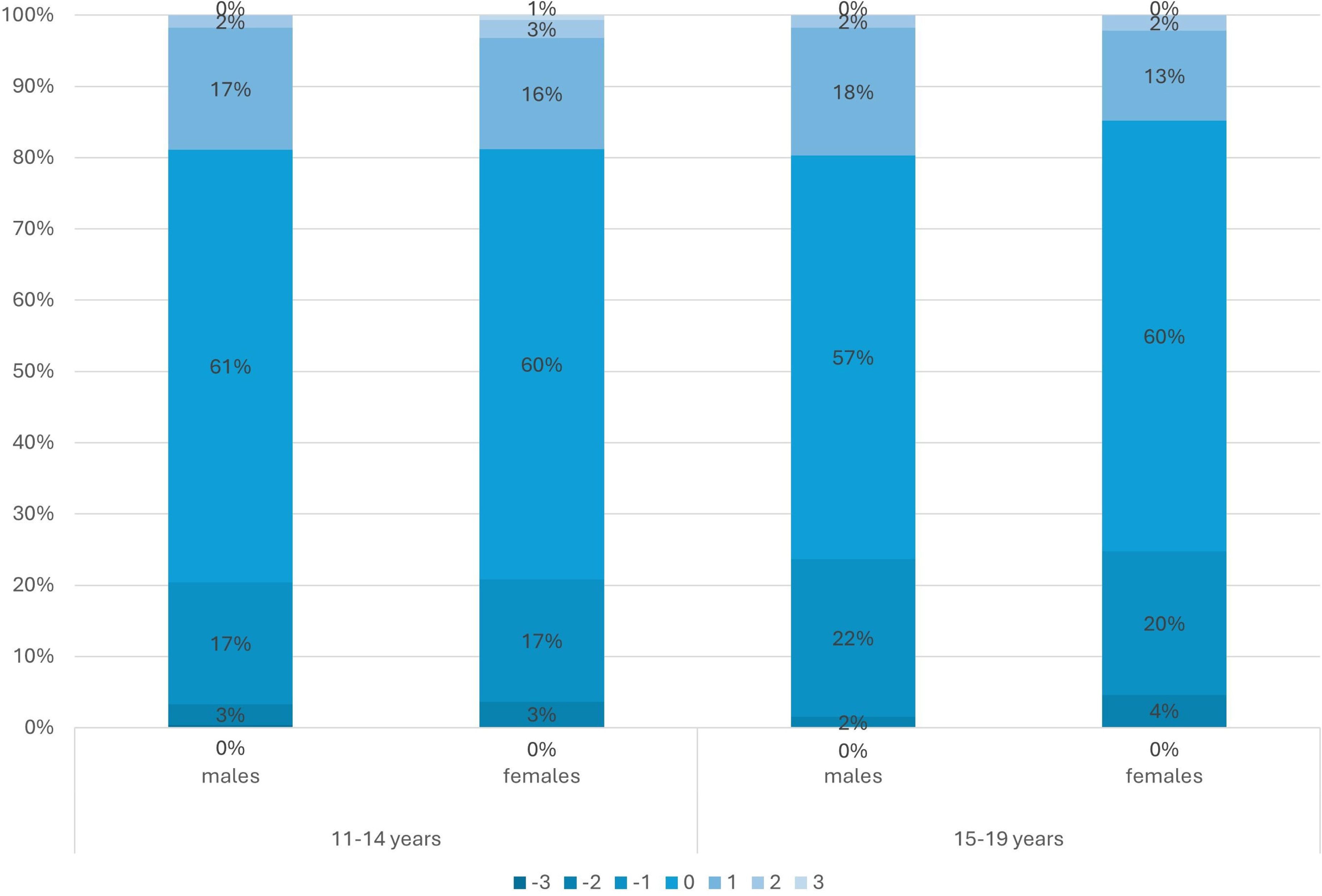
absolute differences (proxy-self) in reported school stress per age group and gender

**Figure 5.**
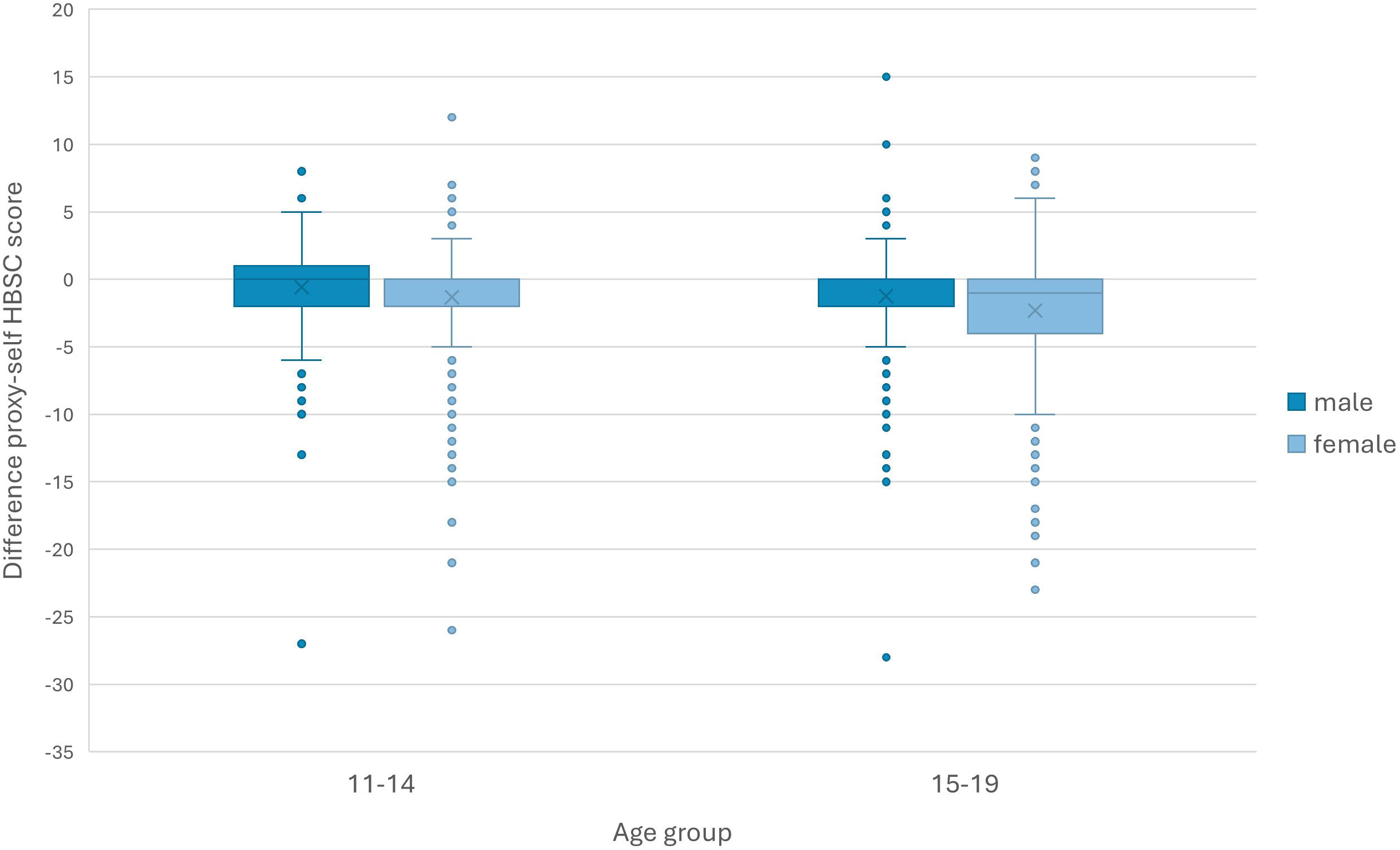
Box plots of absolute differences in proxy and self-reported sum scores of psychosomatic complaints per age group and gender

**Table 3.** Reliability analyses: Pairwise comparisons of proxy- and self-reported answers for school stress and HBSC scum score.

| school stress |  |  |  |  |  |  |
| --- | --- | --- | --- | --- | --- | --- |
|  |  | p-value* | % proxy<self | % proxy>self | Cohen's Kappa [95% ass. CI] | p-value# |
| male | 11-14 | n.s. | 20% | 19% | 0.471 [0.404;0.538] | <0.001 |
|  | 15-19 | n.s. | 24% | 20% | 0.498 [0.433;0.563] | <0.001 |
| female | 11-14 | n.s. | 21% | 19% | 0.445 [0.376;0.514] | <0.001 |
|  | 15-19 | 0.001 | 25% | 15% | 0.509 [0.443;0.574] | <0.001 |
| HBSC sum score |  |  |  |  |  |  |
|  |  | p-value* | Mean (proxy-self) [95% CI] | 5% trimmed mean (proxy-self) | ICC [95% CI] | p-value+ |
| male | 11-14 | 0.001 | -0.59 [-0.87;-0.30] | -0.38 | 0.714 [0.649;0.767] | <0.001 |
|  | 15-19 | <0.001 | -1.22 [-1.62;-0.83] | -0.96 | 0.763 [0.683;0.820] | <0.001 |
| female | 11-14 | <0.001 | -1.34 [-1.72;-0.96] | -1.04 | 0.763 [0.677;0.822] | <0.001 |
|  | 15-19 | <0.001 | -2.33 [-2.82;-1.84] | -1.98 | 0.735 [0.582;0.820] | <0.001 |

*\*p-values refer to paired Wilcoxon Test; # p-values refer to the Null hypothesis Cohen's Kappa=0; + p-values refer to the Null hypothesis: ICC=0*

**Table 4.** Pairwise comparison between proxy and self-reports of lifestyle parameters per age group and gender.

|  |  | physical activity |  |  |  |  |
| --- | --- | --- | --- | --- | --- | --- |
|  |  | p-value paired Wilcoxon test | % proxy<self | % proxy>self | Cohen's Kappa [95% ass. CI] | p-value |
| male | 11-14 | <0.001 | 36.7% | 11.0% | 0.405 [0.336;0.474] | <0.001 |
|  | 15-19 | <0.001 | 30.9% | 11.5% | 0.436 [0.367;0.505] | <0.001 |
| female | 11-14 | <0.001 | 34.8% | 7.9% | 0.383 [0.316;0.450] | <0.001 |
|  | 15-19 | <0.001 | 28.7% | 15.5% | 0.477 [0.406;0.547] | <0.001 |
| parental help with school problems |  |  |  |  |  |  |
|  |  | p-value paired Wilcoxon test | % proxy<self | % proxy>self | Cohen's Kappa [95% ass. CI] | p-value |
| male | 11-14 | 0.026 | 28.4% | 22.5% | 0.529 [0.457;0.600] | <0.001 |
|  | 15-19 | n.s. | 29.6% | 26.3% | 0.469 [0.404;0.535] | <0.001 |
| female | 11-14 | <0.001 | 34.7% | 19.3% | 0.521 [0.461;0.581] | <0.001 |
|  | 15-19 | 0.029 | 26.8% | 22.1% | 0.580 [0.522;0.638] | <0.001 |
| digital media use (school) |  |  |  |  |  |  |
|  |  | p-value paired Wilcoxon test | % proxy<self | % proxy>self | Cohen's Kappa [95% ass. CI] | p-value |
| male | 11-14 | n.s. | 18.2% | 16.2% | 0.648 [0.603;0.693] | <0.001 |
|  | 15-19 | n.s. | 25.2% | 25.7% | 0.678 [0.628;0.727] | <0.001 |
| female | 11-14 | n.s. | 19.7% | 21.5% | 0.625 [0.573;0.677] | <0.001 |
|  | 15-19 | n.s. | 21.8% | 24.6% | 0.657 [0.605;0.709] | <0.001 |
| digital media use (private) |  |  |  |  |  |  |
|  |  | p-value paired Wilcoxon test | % proxy<self | % proxy>self | Cohen's Kappa [95% ass. CI] | p-value |
| male | 11-14 | 0.001 | 25.1% | 13.2% | 0.631 [0.578;0.684] | <0.001 |
|  | 15-19 | 0.001 | 31.2% | 19.9% | 0.504 [0.442;0.567] | <0.001 |
| female | 11-14 | <0.001 | 25.8% | 15.1% | 0.632 [0.580;0.684] | <0.001 |
|  | 15-19 | <0.001 | 30.4% | 14.6% | 0.589 [0.534;0.644] | <0.001 |

### 3.3 A dual perspective: Lifestyle parameters per gender and age group

Parental help with school problems, weekly hours of physical activity, and hours of digital media use for school and private concerns were examined from a dual perspective.

Differences between age groups per gender are reported in the text, while Table 3 shows the differences between proxy- and self-reported data.

Proxy-reported help with school issues differed significantly between age groups for boys (p<0.001) and girls (p<0.001), with higher levels of help in younger participants. Proxy-reported use of digital media for school issues differed significantly between age groups for boys (p<0.001) and girls (p<0.001), with higher levels of use in older participants.

Self-reported parental help with school issues differed between 11-14 aged and 15-19 aged boys (p<0.001) and girls (p<0.001) significantly with less help in the older age group. Hours of digital media use for school concerns differed significantly between 11-14 aged and 15-19 aged boys (p<0.001) and girls (p<0.001) with more hours in the older age group.

Proxy- and self-reported hours of digital media use for private issues differed significantly in the age group from 11-14 for boys and girls and in the age group from 15-19 years for boys and girls, with higher levels of media use in self-reports. No significant differences were found between proxy and self-reports regarding the hours of digital media use for school issues. Proxy- and self-reported help with school problems differed significantly in the age group from 11 to 14 for boys and girls and in the age group from 15 to 19 for girls, with higher levels of help reported by adolescents. Proxy- and self-reported physical activity differed significantly in both age groups for both genders with higher rates of physical activity reported by adolescents.

The corresponding percentages of proxy reports rated lower (proxy<self) and proxy reports rated higher (proxy>self) than self-reports are reported in Table 3.

Cohen’s Kappa compared proxy- and self-reports and showed moderate associations between proxy- and self-reports regarding perceived physical activity and parental help with school problems. Higher correlations were found for the number of hours of digital media use for school and private concerns.

### 3.4. Associations of school stress and psychosomatic complaints with lifestyle parameters

Significant Spearman’s correlation coefficients (after Bonferroni correction for 2*3=6 tests, thus p<0.05/6=0.008 significant) comparing school stress and psychosomatic complaints with lifestyle parameters for proxy and self-reports per gender and age group are shown in Table 5. The highest relationship was found between school stress and parental help with school problems in all age and gender groups in proxy reports. In self-reports, the highest relationship between school stress and parental help was detected in the male age group from 15 to 19. Furthermore, in the female age group from 11 to 14, the highest relationship with school stress was found for digital media use for private concerns. For psychosomatic complaints, in proxy-reports, the highest relations for males were found with parental help with school problems in both age groups and with digital media use for private concerns in the older age group. For girls, proxy reports reported the highest relationships in the younger age group for private media use and in the older age group for help with school problems, physical activity, and private media use. In self-reports, for males in the younger age group, the highest relationship was found with private media use. In the older age group, a relationship was only found for physical activity. For girls, in the younger age group, the highest relationship was clearly with media use for private concerns, and in the older age group, media use for private concerns and physical activity were related to higher psychosomatic complaints.

**Table 5.**
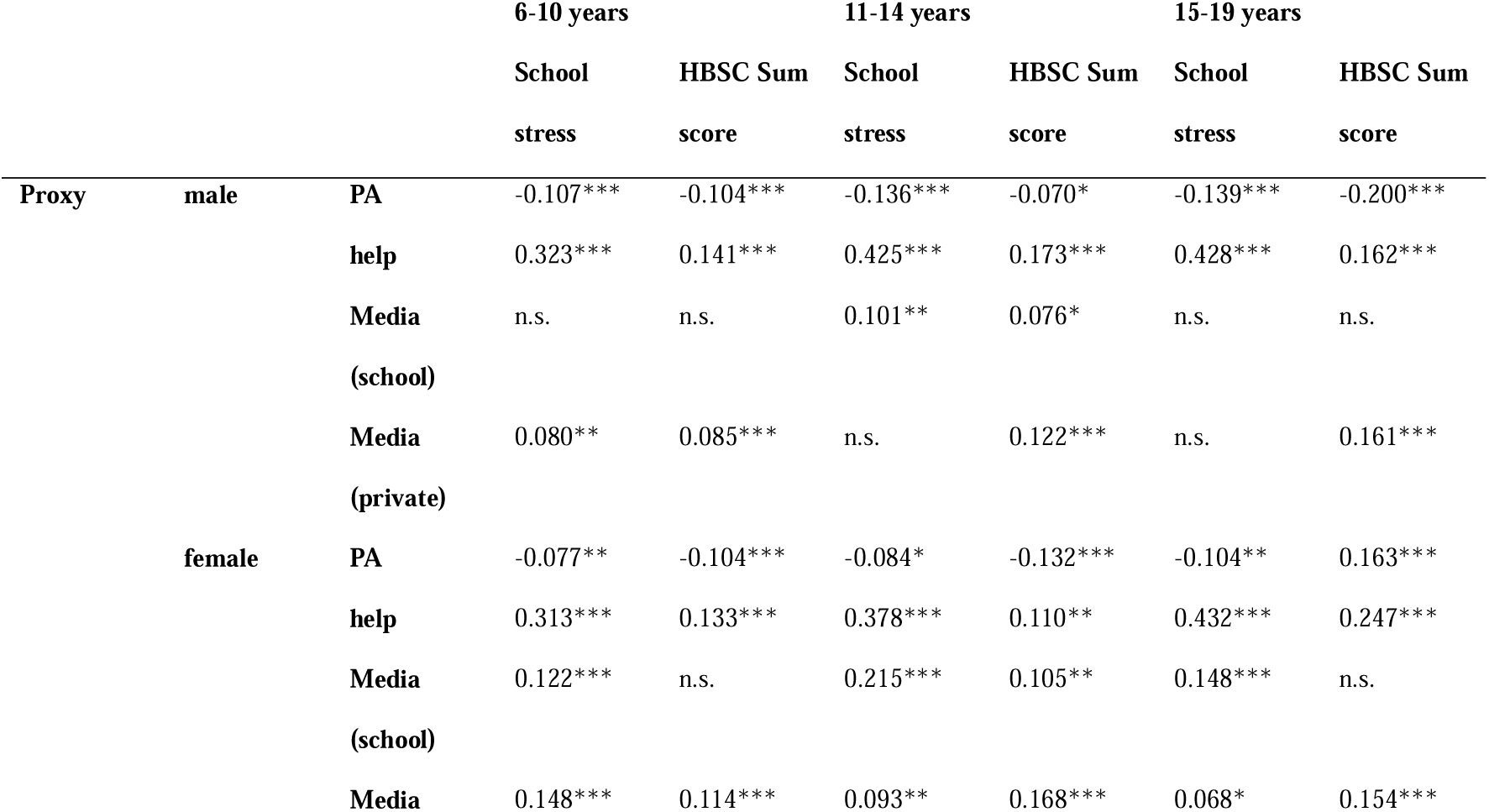
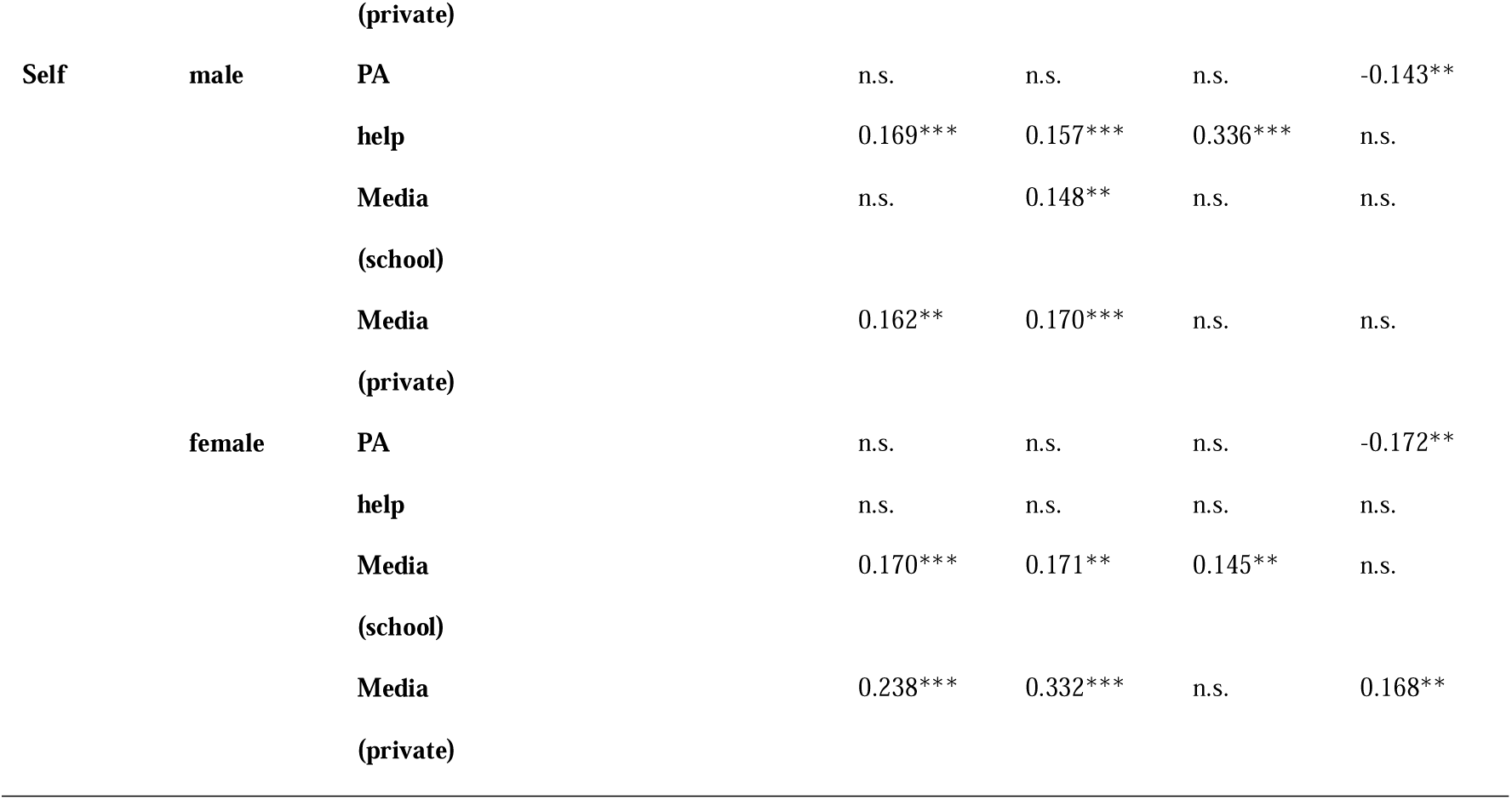
Correlations between sum scores of psychosomatic problems and school stress with lifestyle parameters per age group and gender.

## 4. Discussion

This study analyzed proxy- and self-reported data on school stress and psychosomatic complaints, stratified by age and gender. Higher rates of school stress and psychosomatic complaints were found in girls between 11 and 19 years in proxy- and self-reports, while younger boys between 6 and 10 years exhibited more school stress than girls. Boys in all age groups were more likely to engage in physical activity, and in the oldest age group, they used digital media more often for private concerns, while in this age group, girls were more likely to use digital media for school concerns. Parents perceived their children’s psychosomatic problems as less severe than adolescents did, while there was little difference in perceived school stress between parents and children. Only in the group of girls between 15 and 19 years did parents underestimate their daughters’ school stress. Parents perceived that their children did less physical activity and used digital media less than their children stated in self-reports, while there was no difference between proxy and self-reports for digital media use for school concerns and only a small difference for perceived parental help with school problems. While parents stated that school stress was related especially to help with school problems, in self-reports, parental help received less weight in relation to school stress, while digital media use for private concerns received more weight. While in parent-reports psychosomatic complaints were mainly related to help with school problems and digital media use for private concerns, in self-reports, higher relations were detected for physical activity, digital media use for school and private concerns, and only small relations with parental help with school problems.

### 4.1 Demographic situation

Demographic factors such as age, family background, and gender change with the age of children and adolescents. As children grow older, single parenthood becomes more common, and the migration background tends to be higher among younger children, as well as parental education. These results correspond to the natural development of the population in the area. These factors can affect children’s performance in school and their overall well-being. In (11),(40), the authors find strong relationships between school issues and single parenthood. Children of single parents are one of the most vulnerable subgroups when examining school stress. The results may help primary care providers identify at-risk adolescents and their problems and establish timely prevention and care. School-difficulty prevention should consider family features and include early monitoring of behavior and health-related difficulties in adolescents.

### 4.3 School stress and psychosomatic complaints in different age groups per gender

For children, nearly no gender disparities were found. Additionally, parent-reported school outcomes and psychosomatic complaints were mostly related to sociocultural problems like FAS III, CASMIN index, MSPSS and single parenthood. This result indicates that family support especially is needed in young children to give them a good school and health start for adolescence. In early and late adolescence, gender had the largest relation to all outcomes indicating more stress and more psychosomatic complaints in girls. Results underscore the gender gap discussed in (12),(13),(14),(12). Additional to the increasing gender gap, our results indicate that longitudinal studies are needed to understand the development of the relation between school stress and psychosomatic complaints over years and how the gender-gap starts to develop from different socio-demographic backgrounds in childhood.

Despite the gender gap, especially in proxy reports, we found higher levels of perceived school stress and psychosomatic complaints in single-parent reports and in reports having a low or middle MSPSS. These results align with results from (11), (40), (1), (27), (23), underscoring the importance of focusing in prevention planning on these vulnerable subgroups.

### 4.3 Parental and adolescents perceived school stress and psychosomatic complaints

Psychosomatic complaints were reported more frequently by adolescents than by their parents. Former research (23) reported that child and parent reports can diverge considerably from one another, especially if the behavior or trait is difficult to observe or estimate objectively. Parents and children are more likely to report objectively observable outcomes in the same way. Other studies on depressive symptoms or behavioral problems (23),(24),(25) found higher levels in self-reports than in proxy reports, indicating that relying solely on parents may result in young people with problems being missed and, therefore, untreated. As adolescents’ psychosomatic complaints may lead to higher mental health problems in young adulthood (15),(16),(17),(18),(19), our results mark an important step in monitoring psychosomatic complaints reported by parents and children. To prevent future mental health issues, it is important to rely on adolescents’ self-reported psychosomatic complaints and develop strategies to help them in the early developmental phase.

Late adolescent girls reported more school stress than their parents. Other age groups were not affected by parent-child discordance in school stress. This may indicate that parents underestimate their big daughters’ school burdens. This subgroup can be identified as a vulnerable group to be monitored by parents and teachers for the early detection of school stress. Longitudinal studies are needed to monitor the development of psychosomatic complaints, school stress, and targeted early interventions. Intervention may focus on two main points: training parents in the detection and observation of psychosomatic complaints in their children and providing tools for adolescents to deal with their psychosomatic complaints. Intervention programs should involve parents and children with a special focus on vulnerable subgroups, such as late adolescent girls and single-parent families.

### 4.4 Adolescents’ psychosomatic complaints and stress and relations to lifestyle

School stress was positively related to helping with school issues in all age groups for both genders, but the relationships were much more evident in proxy than in self-reports. In girls, self-reported help with school issues was not or only weakly related to school stress and even less so to psychosomatic problems. This dual perspective shows that parents tend to relate help with school problems to school stress. They may tend to solve school stress by helping with school problems or, conversely, cause school stress by trying to help with school problems. Furthermore, in adolescent girls, parents related school stress to increased engagement with digital media for school issues. Parental perspectives on school stress mainly relate to other school-dependent factors. On the other hand, in self-reports, early adolescents tend to relate school stress to digital media use for both private and school-related concerns. In late adolescence, parental perceptions are closer to adolescents’ perceptions.

Thus, the dual perspective on school stress reveals differences, especially in early adolescence, indicating a better monitoring of private and school concerning digital media use instead of exhaustive help with school issues. This finding, consistent with broader screen time research (7), suggests that for this vulnerable group, strategies should consider not only general screen time reduction but also mindful engagement with digital media, even for school-related tasks. This phenomenon disappears in late adolescence. In (40), (8), the relevance of the school setting in support of prevention among early adolescents has been demonstrated. The authors highlighted that selecting and managing potentially useful information (instead of letting the information load become a hindrance) is something that can be learned. Together with our results, we can identify the vulnerable subgroup of early adolescents when addressing school-based interventions to reduce screen time. For this group, it is not enough to promote sophisticated screen time use in general; school homework and school issues may not focus on digital media use.

For psychosomatic complaints, in children from 6 to 10 years, school played a small role. The results are supported by the aforementioned relations to socioeconomic variables in this age group. In the older age groups, for boys in proxy reports, the results were similar to the children’s reports, with slightly higher links to parental help with school problems, while in self-reports, the only association was found with less physical activity. For early- and late-adolescent girls, in self- and proxy reports, psychosomatic complaints were mainly related to non-school-related lifestyle factors. Parents perceived a relationship between parental help with school problems and only early adolescent girls.

The authors of (27) discussed the direct effects of school pressure and physical activity on psychosomatic complaints. They suggested that supporting young people in managing school demands and promoting their engagement in physical activities could be effective in alleviating psychosomatic complaints. Generally, this indication can be applied to our cohort, but should be extended to reduced digital media use, especially in late adolescent girls.

### 4.5 Methodological considerations

In Northern Italy, as in other high-income countries, school stress is an important correlate of psychosomatic complaints in children and adolescents, and this correlation does not change with age. Conversely, other correlations changed from childhood to late adolescence. While in childhood, the gender gap was not evident, it became important in early and late adolescence. Other demographic factors were related to psychosomatic complaints in childhood (6–10), indicating that early family support in primary schools, especially for families with low education levels and low socioeconomic status and for single parents, may reduce school stress in children. Longitudinal research is needed to understand the development of psychosomatic complaints from socio-demographic effects to gender-related effects.

The gender gap appears during early adolescence. Girls are affected by more psychosomatic complaints and more school stress, and even the use of digital media for school and private issues affects their psychosomatic well-being. Even if boys report significantly fewer psychosomatic complaints, these related factors are the same for both genders. Sophisticated digital media strategies, even in school settings, may help reduce psychosomatic problems in this age group. School-based intervention programs to teach early adolescents how to use digital media in school and private contexts can be introduced, but further research is needed to understand how they may affect health-related parameters.

Parent-child disagreement in reporting was evident in our study, particularly for school stress and psychosomatic complaints, with higher levels of perceived problems in self-reports. This highlights the importance of using self-reports when assessing youth health outcomes. Parent reports can provide further indications and can be used as supportive information but can never substitute self-reported outcomes.

Finally, school stress and psychosomatic complaints were associated with lower physical activity. The theme has been discussed in the literature (3),(41),(7), proposing public health strategies to decrease screen time and increase physical activity to lower psychosomatic complaints among adolescents from high-income countries. Consequently, in Northern Italian children and adolescents, school stress remains a major problem when discussing psychosomatic complaints. Physical activity can reduce these complaints, but independently of perceived school stress. Many studies mention both more physical activity and less screen time use when proposing how to achieve a better quality of life.

### 4.6 Strength and Limitations

This study had several strengths. This study investigates the increasing problem of school stress and psychosomatic complaints in high-income countries in Northern Italy, where evidence is lacking. The findings highlight the different needs of different age groups, the associations between digital media use and physical activity for school issues, and specific aspects of the gender gap. These aspects underscore the need for differentiated longitudinal research in this region and other high-income countries. Data were derived from a large population-based survey representing the population of South Tyrol. Double proxy self-reported data allow for comparison and identification of discrepancies between parental and child-perceived health and stress parameters. This dual perspective has been found to be important in mental health research on young people. Our study extends the research to investigate the earlier stages of psychosomatic complaints. These new insights may contribute to prevention planning.

The cross-sectional design of this study limits our ability to infer causality, allowing us to identify only associations between variables. Longitudinal studies are therefore needed to understand the developmental trajectories of school stress from childhood to late adolescence and to obtain more detailed information about the gender gap.

Finally, our anonymous online survey yielded a response rate of approximately 23%. The value and practicability of online child mental health surveys were discussed in (42), showing that with a response rate of approximately 20%, results mostly replicate results from other studies. For pandemic times, the usability of online surveys was discussed in (43), stating that with email invitation, a percentage of about 30% is reachable and is comparable to other surveys. In our study the age and gender of the schoolchildren corresponded to official statistics as well as the percentage of single parents. Nonetheless, there is a potential for selection bias (e.g., higher educated families are more likely to participate) and reporting bias (e.g., social desirability in responses), which may have influenced the observed prevalence and associations.

## 4. Conclusions

This population-based dual-informant study reveals that the association between school stress and psychosomatic complaints is consistent across all age groups and both genders, but that the prevalences as well as the broader pattern of risk factors shift substantially from childhood to late adolescence.

In childhood (ages 6–10), psychosomatic complaints were primarily associated with sociodemographic disadvantages. Therefore, early prevention should prioritize support for socioeconomically disadvantaged and single-parent families during primary school years.

The gender gap emerges during early adolescence. The findings confirm, for the first time in an Italian regional cohort, the gender-stratified patterns documented in the Northern European HBSC studies.

Digital media use and its associations with school stress and psychosomatic complaints need to be monitored, specifically in early adolescents of both genders. Physical activity was associated with fewer complaints, particularly among girls. Parent–child reporting discrepancies were evident throughout, highlighting the importance of systematically including adolescent self-reports in health surveillance. Parental help with school problems is important but should not substitute the importance of guided and sophisticated digital media use.

The cross-sectional design limits inferences to associations, and the developmental shifts observed across age groups require confirmation through longitudinal follow-up within the COP-S survey series. Nevertheless, the present findings identify actionable prevention targets: family support in primary school, gender-sensitive monitoring of school stress from early adolescence onward, guided digital media use for school tasks, and systematic inclusion of adolescent self-reports in health surveillance.

## List of Abbreviations

CASMIN: Comparative Analysis of Social Mobility in Industrial Nations
CI: Confidence Intervals
COP-S: Corona and Psyche South Tyrol
FAS: Family Affluence Scale
HBSC-SCL: Health Behavior in School-aged Children Symptom
CheckList ICC: IntraCorrelation Coefficient
MSPSS: Multidimensional Scale of Perceived Social Support
n.s.: Not significant
VIF: Variance Inflation Factor

## Statements and Declarations

### Data availability

The datasets generated and analyzed during the current study are available from the corresponding author upon reasonable request.

### Ethical approval

The study was conducted in accordance with the Declaration of Helsinki and approved by the Ethics Committee of the Autonomous Province of Bolzano, Italy (protocol code 11-2025, 19 February 2025).

### Informed consent

Written informed consent was obtained from all participating parents/guardians and adolescents.

### Authors’ contributions

Conceptualization: V.B., and C.J.W.; Investigation: V.B.; Analysis: V.B.; Writing—original draft: V.B.; Writing—review and editing: C.J.W., G.P. and A.E.; Supervision: A.E., D.H. and G.P.

## Acknowledgments

The authors thank the school authorities and the management of public schools in South Tyrol for facilitating recruitment and participation. The authors also thank Irene Parnigotto for revising the Italian version of the questionnaire.

