## Supplementary Table 1 for "Linking School Stress and Psychosomatic Complaints in South Tyrol, Northern Italy: Parental and adolescents’ perspectives in a cross-sectional design"

1 S1 Detailed percentages for school stress and mean+-SD for the sum score of psychosomatic complaints per gender, age group and informant type

|  |  | 6-10 y | 11-14 y |  | 15-19 y |  |
| --- | --- | --- | --- | --- | --- | --- |
|  |  | proxy | proxy | self | proxy | self |
| school stress | | % | % | % | % | % |
| males | not at all | 29% | 14% | 13% | 10% | 11% |
|  | a little | 50% | 53% | 53% | 51% | 46% |
|  | rather | 16% | 23% | 23% | 25% | 29% |
|  | very much | 5% | 10% | 10% | 14% | 14% |
| females | not at all | 30% | 14% | 16% | 7% | 4% |
|  | a little | 53% | 53% | 54% | 42% | 38% |
|  | rather | 13% | 22% | 23% | 31% | 34% |
|  | very much | 3% | 12% | 8% | 19% | 23% |
| Sum score of psychosomatic complaints | | M+-SD | M+-SD | M+-SD | M+-SD | M+-SD |
| males |  | 11.04+-3.06 | 10.95+-2.82 | 11.52+-3.50 | 11.55+-3.93 | 12.86+-4.65 |
| females |  | 10.94+-2.85 | 12.03+-3.79 | 13.42+-5.11 | 12.94+-4.66 | 15.23+-6.30 |
